# Approximate relationships between SIR and logistic models

**DOI:** 10.1101/2020.07.08.20149070

**Authors:** David E. Clark, Gavin Welch, Jordan S. Peck

## Abstract

Infectious epidemics are often described using a three-compartment Susceptible-Infectious-Removed (SIR) model, whose solution can be shown to involve generalizations of the logistic distribution. Using mathematical relationships relating these generalized logistic distributions, the population proportion remaining Susceptible can be approximated using the inverse of a standard cumulative logistic distribution, while the population proportion actively Infectious can be approximated using the density of a logistic or log-logistic distribution. Conversely, the parameters of an underlying SIR model can be approximately inferred from population-based data that have been estimated using logistic and/or log-logistic models.

## INTRODUCTION

Models of infectious epidemics in a population have often been based on a system of differential equations describing the theoretical transition of individuals through a series of states from Susceptible (S) to Infectious (I) to Removed (R). Recent experience has also shown value in fitting emerging infection data to a logistic growth curve for the purpose of short-term planning. The aim of this study is to show how assumptions and analyses using one of these approaches may be approximately transferred to the other.

The SIR model, attributed to Kermack and McKendrick (1927), is a three-compartment model where individuals transition from Susceptible to Infectious at a rate proportional to the product of the number of individuals in these two compartments, and then move from Infectious to Removed at a rate proportional to the number of individuals in the Infected compartment.

This may be expressed as the system of differential equations

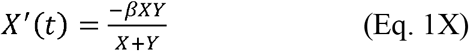

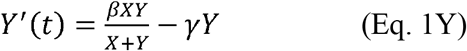

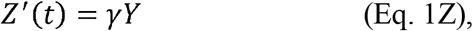

where *X* is the number Susceptible, *Y* is the number Infectious, and *Z* is the number Removed.

Bohner and colleagues (2019) have shown that a solution to the SIR system described above is given by

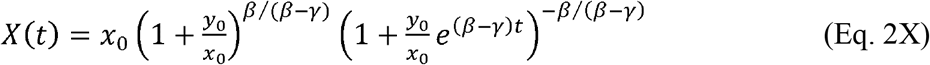

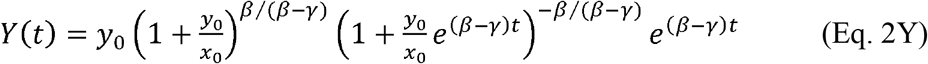

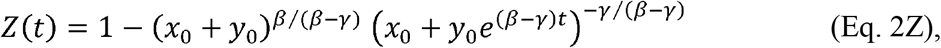

subject to initial conditions *x*_0_ and *y*_0_, with *x*_0_ + *y*_0_ ≤ 1.

Using the substitutions

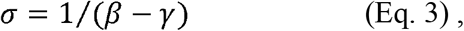

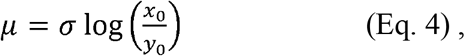

and

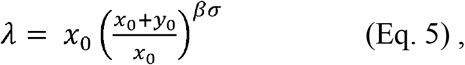

Equations 2X, 2Y, and 2Z simplify to

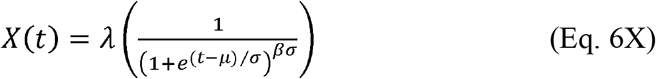

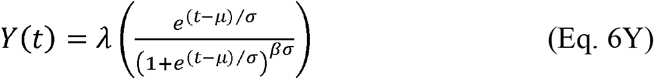

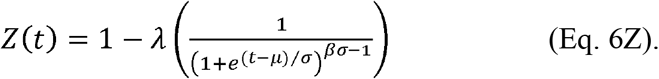

It is apparent that these results are affected not only by the ratio *β* /*γ* (sometimes called *R*_0_), but also by the difference *β* −*γ* ; the quantity *βσ* is equivalent to *R*_0_ / (*R*_0_-1). It is also apparent that there is some resemblance to a logistic model.

## APPROXIMATING *X*(*t*) WITH AN INVERSE LOGISTIC CURVE

With the substitution *θ* = *βσ*, Equation 6X may be rewritten

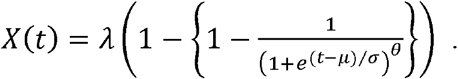

The bracketed term is the Cumulative Distribution Function (CDF) of a “Type 2” Generalized Logistic Distribution (Johnson *et al*, 1994), which may be abbreviated GL2 and is sometimes also called a “Reversed Generalized Logistic Distribution” (El-Saidi *et al*, 1996). It is a special case of the Richards Model (Richards, 1959), which has been widely used in biology and population studies. The GL2 Distribution is similar to the Logistic Distribution, and is identical when *θ* = 1. *X*(*t*) involves the inverse of a GL2 Distribution, so a reasonable approximation might be expected using the inverse of a Logistic Distribution, which may be written

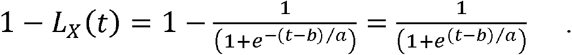

The mean, variance, and other moments of a GL2 Distribution can be calculated in terms of the Gamma Function and its derivatives, and demonstrate that it will be negatively skewed (longer tail to the left) if *θ* > 1 and positively skewed if *θ* < 1 (Dubey, 1969; El-Saidi *et al, 1996*). However, it is easier to work with the median and other quantiles of the distribution, which do not require special functions.

Any quantile *q* of a GL2 Distribution can be obtained by inverting its CDF, that is, by equating the CDF to *q* and solving for *t*, which results in

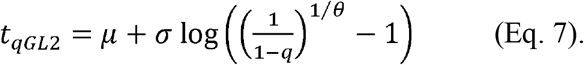

The quantiles for a Logistic Distribution can be obtained from the same equation, letting *θ* =1. Since *X*(*t*) is a linear transformation of the inverse of a GL2 Distribution, the quantile *q* for *X*(*t*) will equal the quantile 1-*q* for the underlying GL2 distribution. The parameters of the SIR model can therefore be used along with Equation 7 to approximate *X*(*t*) with the inverse of a Logistic Distribution. One useful point of reference is the median, which is obtained when *q* = .5, and any other quantile can be used along with the median to estimate parameters for a logistic curve.

A Logistic Distribution will be symmetric with mean, mode, and median all equal to *b*. Equating the median and .75 quantiles for a Logistic Distribution to those for a GL2 Distribution with *θ* =*βσ* results in the approximate relationships

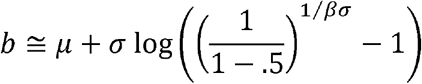

and

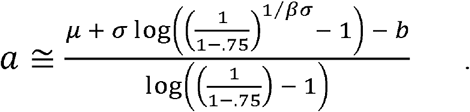

If *x*_0_ is much less than 1, the .75 quantile of the GL2 Distribution may be less than 0, in which case the Logistic Distribution may instead be fitted to the median and the 1-*x*_0_/*λ* quantile, where *t*=0. In either case, if the parameters of an SIR model are known, parameters *a* and *b* can be derived from these relationships to create a logistic curve *λL*_*X*_(*t*) whose inverse 1-*λL*_*X*_(*t*) will approximate *X*(*t*).

## APPROXIMATING *Y*(*t*) WITH A LOGISTIC OR LOG-LOGISTIC CURVE

The quantity 1 - *X*(*t*), or a similar logistic curve, describes the proportion of the population that has acquired the disease. This is valuable from the public health perspective, and is often the focus of attention. However, *Y*(*t*) is also important because it reflects the proportion of the population actively suffering from the disease at a given time, and may therefore be valuable for short-term planning within a health care delivery system.

With the substitution *θ* = *βσ* – 1, Equation 6Y can be rewritten

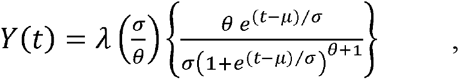

where the expression inside the brackets is the Probability Density Function (PDF) for another GL2 Distribution. By equating the median and another convenient quantile as described above, the parameters for another Logistic Distribution can be derived, whose PDF may be written

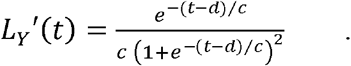

An approximation for Y(t) is then obtained by multiplying *L*_*Y*_ ′(*t*) by the quantity *λσ* /*θ*, which is also equal to *λ* / *γ*.

If *βσ* – 1 is much less than 1, the GL2 Distribution describing *Y*(*t*) will be too skewed to be approximated well by a symmetric Logistic Distribution. However, Log-Logistic (LL) Distributions are similarly skewed, and can serve this purpose. A LL random variable may be defined as one whose logarithm has a Logistic Distribution. LL Distributions have been used for many areas of time-to-event analysis; they are similar to Log-Normal Distributions, but have more convenient mathematical properties (Clark and El-Taha, 2015).

The PDF of a standard two-parameter LL Distribution is constrained to be 0 when *t* = 0, so to allow y_0_ > 0 it is necessary to employ a three-parameter or “shifted” LL Distribution (Ahmad *et al*, 1988), which may be abbreviated LL3. An LL3 random variable has an additional parameter (here represented by *k*), resulting in CDF

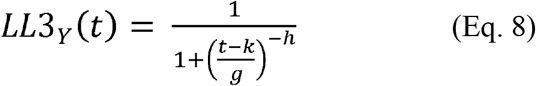

and PDF

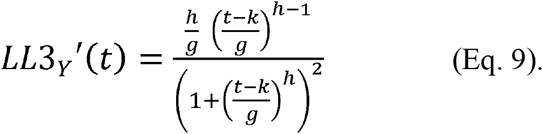

Quantiles of an LL3 distribution can be obtained by inverting Equation 8 to get

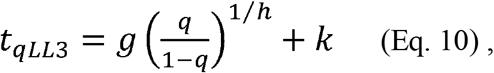

which has the convenient result that the median is simply *g+k*.

*Y*(*t*) may be approximated with an LL3 Distribution by first considering *k* to be a sufficiently small quantile (e.g., 0.005) of *Y*(*t*). Using this value of *k* along with the median and 0.75 quantiles, it is possible to derive

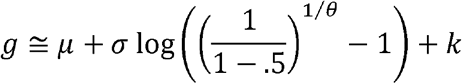

and

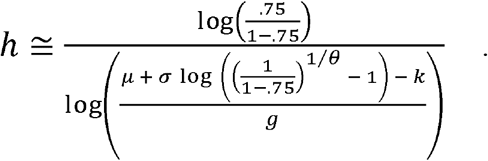

If the .75 quantile of the GL2 Distribution is less than 0, then the LL3 Distribution may instead be fitted to the median and the 1 - (*x*_0_+*y*_0_) /*λ* quantile, where *t*=0. In either case, *Y*(*t*) may then be estimated as *λσ* /*θ* * *LL3*_*Y*_ ′(*t*).

## APPROXIMATING *Z*(*t*) WITH A LOGISTIC OR LOG-LOGISTIC CURVE

Equation 6Z can be rewritten

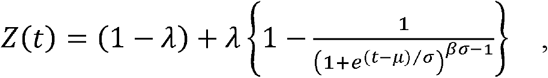

where the bracketed term is the CDF of a GL2 Distribution with *θ* = *βσ* -1. Thus, if *Y*(*t*) has been approximated using the PDF of a logistic or three-parameter log-logistic function, *Z*(*t*) can be approximated using the CDF of the same function, that is, either (1-*λ*) + *λ L*_*Y*_ (*t*) or (1-*λ*) + *λ LL3*_*Y*_ (*t*). This makes sense from the SIR model, since all subjects pass through the Infectious compartment and are then absorbed by the Removed compartment.

## ESTIMATING THE SIR MODEL FROM POPULATION DATA

Once initial conditions *x*_0_ and *y*_0_ have been specified, the SIR model depends only upon the parameters *β* and *γ*, but these are difficult to separate if limited to the equations described above. Fortunately, a brief consideration of the Digamma Function will produce a useful result. The Digamma Function, often abbreviated Ψ, is defined as the first derivative of the logarithm of the more familiar Gamma Function. It has many interesting properties, but it suffices here to know that

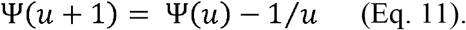

Previous authors (Dubey, 1969; El-Saidi *et al*, 1996) have shown that the mean of a GL2 Distribution is

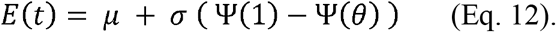

Combining Equations 11 and 12, the difference between the means of two GL2 Distributions with the same values of *μ* and *σ*, one of which has *θ* = *βσ* and the other of which has *θ* = *βσ* -1 is simply

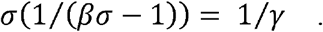

Therefore, if an inverse logistic estimate with parameters *a* and *b* has been made for *X*(*t*) and a logistic estimate with parameters *c* and *d* or an LL3 estimate with parameters *g, h*, and *k* has been made for *Y*(*t*), the difference in their means can be used to estimate *γ*, that is, either

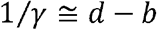

or

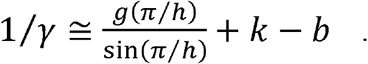

Other sources of information may also provide an estimate for 1/*γ* (which may be interpreted as the mean duration of the Infectious state). Once *γ* has been estimated, *β* can be estimated numerically by determining the value of *β* that minimizes the difference between the approximated median (for either *X* or *Y*) and the median of the corresponding GL2 distribution (Equation 7). Combining Equations 6X and 6Y also shows that when *t* =*μ, X*(*t*) = *Y*(*t*), so *β* can be derived by first estimating *μ* (where the difference between the approximations for *X* and *Y* is minimized) and then solving for *β* using Equations 3 and 4.

Thus, if the observed data for a population undergoing an epidemic can be summarized by a logistic equation for the Susceptible proportion and a logistic or LL3 equation for the Infectious proportion, then the parameters of an underlying SIR model can be inferred. Logistic or LL curves can be estimated from observed data by least squares, maximum likelihood, or other methods. In particular, the LL distribution is a standard option for most statistical software programs used in time-to-event analysis.

To illustrate how models based upon observed population data can be interchanged with the SIR model, Figure 1 shows the exact solutions for a specified compartmental model, corresponding logistic or log-logistic approximations, and a reconstitution of the SIR model derived from the parameters of the logistic or log-logistic approximations. Since the approximations are valid for any specification of *x*_0_ and *y*_0_, piecewise analyses are also possible, for example if some public health or medical intervention may have changed the effective values of *β* and/or *γ*. Figure 2 demonstrates how the values of *X*(*i*) and *Y*(*i*) at some point *t* = *i* can be treated as a new *x*_0_ and *y*_0_ to start a revised model for the next time period.

**Figure 1:**
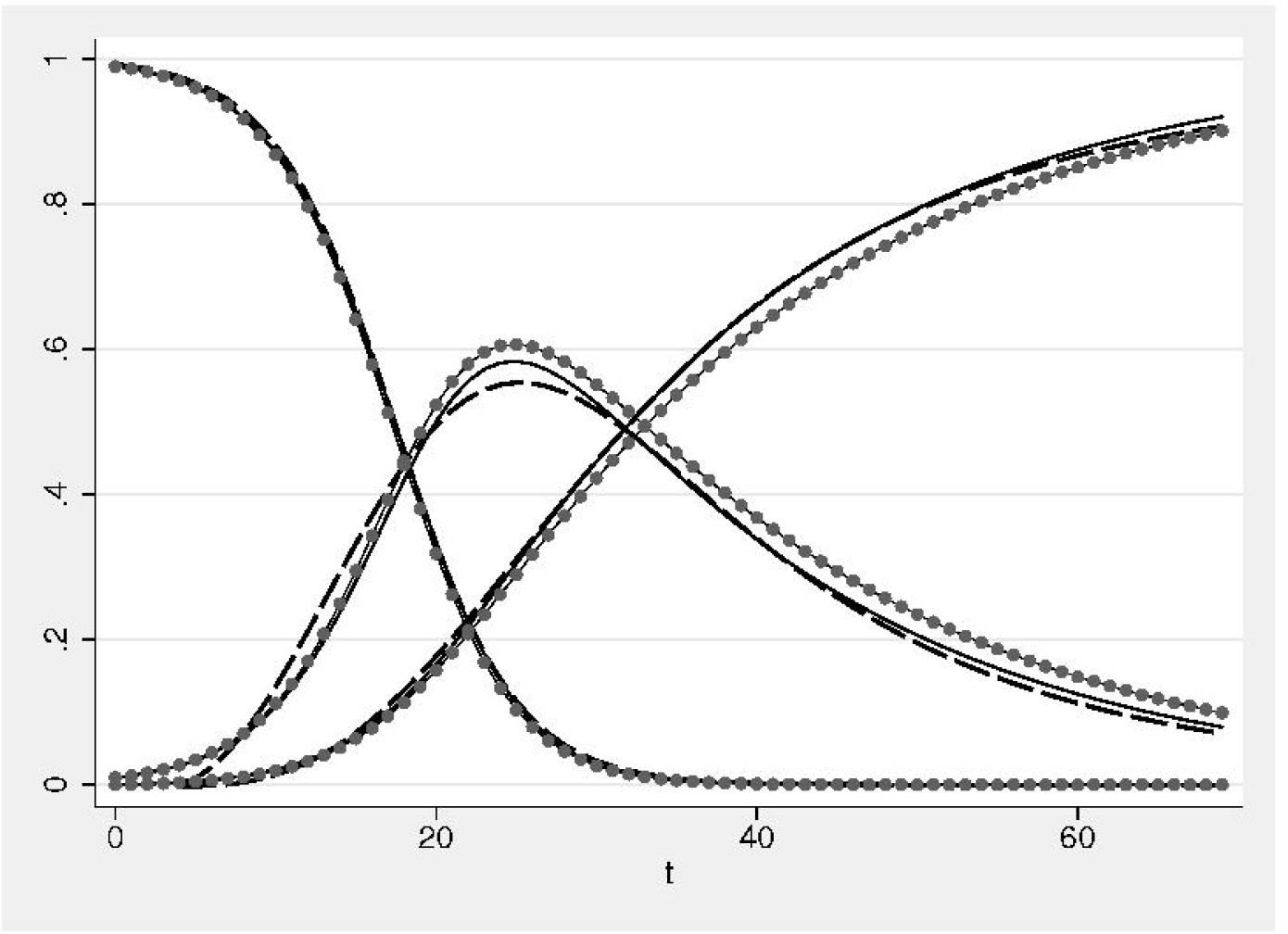
Comparison of SIR model solution (solid lines) with derived approximations (dashed lines) and reconstituted results (dotted lines) for initially specified *x*_0_ = .99, *y*_0_ = .01, *β* =.30, and *γ* =.05.

**Figure 2:**
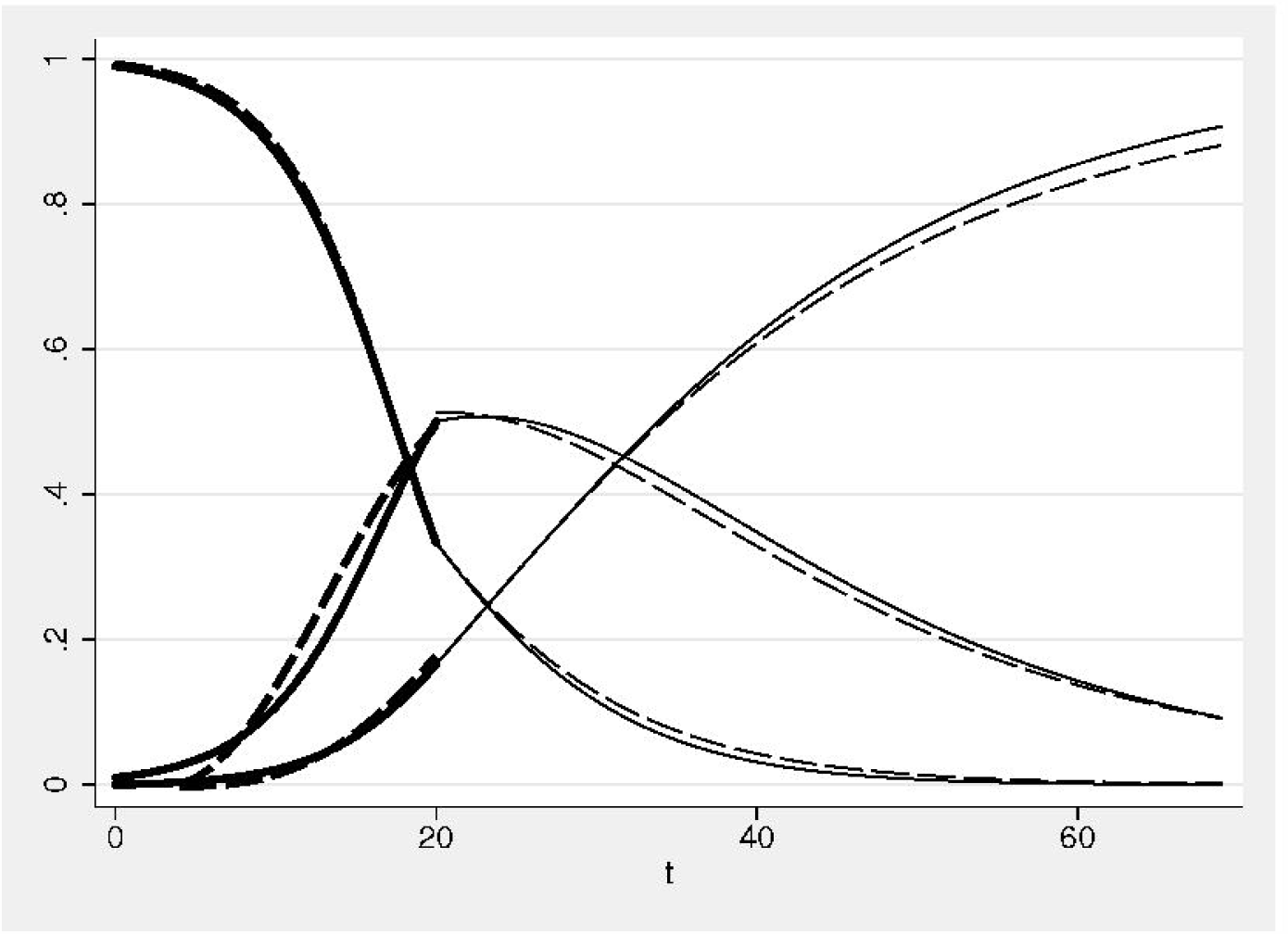
SIR model solutions (solid lines) and approximations (dashed lines) for initially specified *x*_0_ = .99, *y*_0_ = .01, *β* =.30, and *γ* =.05, changing at *t* = *i* = 20 to *x*_*i*_ = .33, *y*_*i*_ = .50, *β* =.15, and *γ* =.05.

## DISCUSSION

Using a logistic curve to approximate *X*(*t*) is obviously inexact, and will be closest when *βσ* is close to 1, that is when *β* /*γ* is large. The logistic approximations for *Y*(*t*) and *Z*(*t*) are also inexact, and will be closest when *βσ* -1 is close to 1, that is when *β* /*γ* is close to 2. The log-logistic approximations for *Y*(*t*) and *Z*(*t*) will generally be preferable when *βσ* -1 is less than about 0.5, that is when *β* /*γ* is greater than 3. The quantiles used for curve-fitting may also need to be adjusted when working with the tails of the distributions. However, given the uncertainty underlying any model, these approximations may be close enough to be useful. If not, the realization that the SIR model can be solved in terms of a GL2 Distribution may itself be of some value.

The ability to easily relate an SIR model to a model based on probability distributions may have direct value to communities responding to an acute epidemic. The facility with which an epidemic can be understood and modeled directly influences how quickly local interventions can be designed and implemented. Epidemic modelers typically rely on labor-intensive contact tracing and patient testing to approximate the key variables to inform an SIR model. This work must often be repeated in different geographical areas as the parameters for a particular infectious agent may differ depending on the nature of the community in which it is spreading. Most epidemiologists are more familiar with statistical distributions than with differential equation modeling, which may limit their ability to utilize the insights of an SIR model.

In any serious epidemic (including seasonal influenza) cumulative infection and symptomatic tracing data are published periodically. Those who are seeking to build models for specific purposes (such as hospital supply planning) can fit the available data to logistic or log-logistic curves using commonly available statistical software, to estimate parameters and rapidly build their own informative models. Being able to relate these findings to the more sophisticated class of SIR models using the simple methods described here may increase confidence in any predictions and improve understanding of the underlying dynamics of disease spread and resource requirements.

## Data Availability

N/A

